# Analyzing the determinants for using health research evidence in health planning in Tanzania: a cross-sectional study

**DOI:** 10.1101/2024.12.16.24319091

**Authors:** Pius Kagoma, Richard Mongi, Albino Kalolo

**Author notes:** Corresponding author: Pius Kagoma; P.O. Box 1923 Dodoma, Tanzania. Richard Mongi; P.O. Box 259, Dodoma, Tanzania, Albino Kalolo; P.O. Box 175, Ifakara, Morogoro, Tanzania.

## Abstract

**Introduction:** Achieving Universal Health Coverage (UHC) requires utilizing research evidence to inform the decision-making process. However, little information is available on the determinants for using research evidence in planning in Lower Middle-Income Countries (LMICs), including Tanzania. This paper aims to investigate the determinants of using health research evidence in health planning in Tanzania.

**Materials and methods:** This study employed a cross-sectional study design. Data on health research evidence and its determinants were collected using a structured questionnaire from 422 respondents from 9 regions of Tanzania from October to December 2023. The data were analyzed using STATA version 18 for descriptive and inferential statistics. The association between variables was determined using a chi-square test at a 95% confidence level.

**Results:** The study revealed that 66.2% of participants strongly agreed to use health research evidence during planning. However, significant barriers were identified, including lack of dissemination (74.5%), inadequate human and non-human resources (70.0%), and insufficient knowledge and training in research (63.7%). A chi-square test confirmed significant associations between these barriers and the reduced use of research evidence (p<0.05). Conversely, more than 70% of respondents identified opportunities such as the availability of research coordinators, university partnerships, available research budgets, and internet access, all significantly associated with increased health research evidence use. More than 50% of participants reported motivational factors that like continuous quality improvement agenda in the healthcare sector, availability of short and long-term courses, on-the-job training, and provision of incentives like extra duty allowances were positively linked to research utilization.

**Conclusion:** The study found that 66.2% of participants used health research evidence in planning, but barriers like lack of dissemination, resource shortages, and inadequate training persisted. Interventions should focus on improving dissemination, resources, and training. Future research should explore strategies for enhancing these interventions.

## INTRODUCTION

Health research evidence plays an essential role in shaping effective health policies and interventions, which are crucial for improving public health outcomes (1). In resource-limited settings like Tanzania, where health challenges are complex and resources are constrained, the strategic use of research evidence is particularly important. It can guide decision-makers in identifying priority areas, optimizing resource allocation, and implementing interventions that are both cost-effective and impactful. Moreover, the use of evidence can help health planning teams avoid repeating past failures and introduce innovative solutions (2). Despite its recognized importance, the extent to which health research evidence is utilized by health planning teams in Tanzania remains unclear. Understanding the factors that influence this utilization is essential for promoting evidence-based decision-making, which is critical for achieving the ambitious health targets set forth in the Sustainable Development Goals (SDGs), particularly Universal Health Coverage (UHC)(3).

Efforts to promote the integration of research evidence into health planning and decision-making have been made in various settings. These include the establishment of knowledge translation platforms, research dissemination strategies, health conferences, and capacity-building initiatives targeting health planning teams. However, the success of these efforts varies significantly across contexts, largely depending on factors such as institutional support, the capacity of the health workforce, and access to reliable data. In Tanzania, while some initiatives aim to bridge the gap between research and practice, the actual impact of these efforts on health planning remains under-explored.

Research across different settings has demonstrated that the use of health research evidence in decision-making is influenced by a variety of factors. These include organizational culture, the availability of resources, the expertise of the planning teams, and access to research outputs [4]. In a study conducted by Kagoma et al (4), it was found that health planning teams in Tanzania consist of professionals from various disciplines, including Administrators (9%), Medical doctors (25.36%), Nurses (23.46%), Laboratory scientists (8.53%), Pharmacists (6.64%),

Radiographers (1.66%), Environmental health officers (3.08%), Nutrition officers (3.08%), Social welfare officers (4.74%), and others (14.46%). Their primary responsibility is to plan, implement, and evaluate health interventions [4]. However, several factors may limit their ability to fully integrate research evidence into their work (5). These include inadequate dissemination of research findings, lack of access to research evidence, limited skills in interpreting research data, and insufficient organizational support (3,6–8).

While previous studies have explored various factors influencing the use of health research evidence in policy and planning, there remains a gap in understanding the specific determinants that affect the utilization of health research evidence among health planning teams in Tanzania(4,9). This study seeks to fill this gap by analyzing the key factors that influence evidence use among these teams at both the regional and council levels, guided by the COM-B Model (Capability, Opportunity, and Motivation) as the theoretical framework (10), which has been widely used in understanding evidence-based decision-making processes. The COM-B model is particularly suited to the Tanzanian context because it focuses on the essential components of behavior change Capability, Opportunity, and Motivation which are critical for understanding and addressing the factors influencing the use of health research evidence by planning teams. By examining these elements, the model helps identify specific barriers and enablers that can be targeted to improve evidence-based decision-making in Tanzania’s resource-limited. By applying this theoretical lens, the study aims to provide a deeper understanding of the barriers and facilitators to using research evidence in health planning, with the following specific objectives: 1) Analyze the current usage of health research evidence among planning team members at the regional and council levels, 2) Analyze the capacity of health planning team members to utilize health research evidence, 3) Identify opportunities for enhancing the use of health research evidence in health planning, 4) Identify opportunities for enhancing the use of health research evidence in health planning, 5) Explore the motivations for using health research evidence among health planning members at both regional and council levels. The findings of this study are expected to contribute to the development of strategies that enhance the integration of research evidence into health planning processes, ultimately leading to more effective health interventions and improved health outcomes in Tanzania.

### Conceptual framework

The conceptual framework for this study was developed after several consultative meetings with different stakeholders and researchers. The theory was adopted and modified from the (Capability, Opportunity, and Motivation) COM-B model. The COM-B Model at the center of a proposed framework is a behavior system involving three essential conditions: Capability, Opportunity, and Motivation, what we term the ‘COM-B system(10,11). This study investigated how the capability, opportunity, and motivation determinants can influence the behavior of the use of health research evidence during health planning as shown in Figure 1

**Figure 1:**
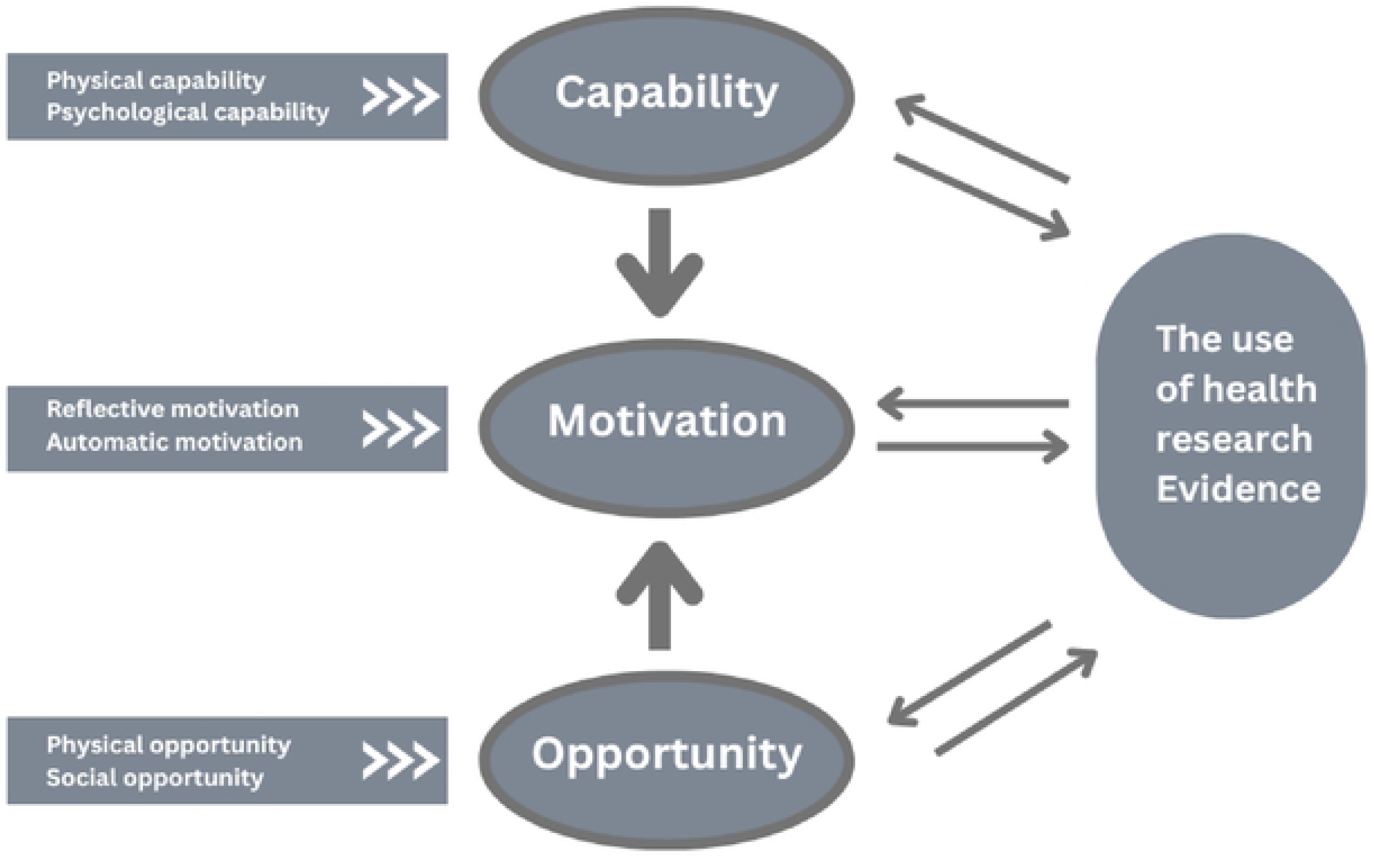
The domains of the COM-B Model modified from Michie et al,.2011(10)

## Materials and Methods

### Study setting

This study was conducted between October and December 2023 in the United Republic of Tanzania, a Lower and Middle low-income country located in East Africa, with a population of about 62 million people. The allocated budget for the country’s Ministry of Health in 2023/2024 was estimated to be 443.6 million US dollars, with 1.2 million US dollars (0.27%) allocated for evidence production. The health research evidence users in Tanzania involve three important Ministries, namely; the Ministry of Health (MoH), The President’s Office Regional Administration and Local Government (PO-RALG), and Health management. The health evidence producers in Tanzania are the National Institute for Medical Research (NIMR), the Tanzania Commission for Science and Technology (COSTECH), Public and Private Universities, Health related institutions or authorities, local and international non-governmental organizations (NGOs), and civil society organizations as shown in Table 1.

**Table 1:** A list of health research evidence producers and users in Tanzania.

| SN | Evidence users |
| --- | --- |
| 1 | Ministry of Health |
| 2 | President's Office Regional Administration and Local Government |
| 3 | Ministry of Education Science and Technology (MEST) |
| 4 | Prime Minister's Office |
| 5 | Regional Health Management Teams |
| 6 | Council Health Management Teams |
| 7 | Healthcare workers |
| 8 | Non-Governmental Organizations (NGOs) |
| 9 | Civil Society Organizations (CSOs) |
| 10 | Private Sector |
| **Evidence producers** |  |
| 1 | National Institute for Medical Research (NIMR) |
| 2 | Tanzania Commission for Science and Technology (COSTECH) |
| 3 | Public Universities; The Muhimbili University of Health and Allied Sciences (MUHAS), University of Dar es Salaam (UDSM), University of Dodoma (UDOM) |
| 4 | Private Universities; Catholic University of Health and Allied Sciences (CUHAS), Kilimanjaro Christian Medical Centre (KCMC), Hubert Kairuki Memorial University (HKMU), St. John's University of Tanzania (SJUT), St. Augustine University of Tanzania (SAUT), Kampala International University (KIU), St. Francis University College of Health and Allied Sciences (SFUCHAS) |
| 5 | Kilimanjaro Clinical Research Institute (KCRI) |
| 6 | Ifakara Health Institute (IHI) |
| 7 | Non-Governmental Organizations (NGOs) |
| 8 | Civil society organizations (CSOs) |
| 9 | Private sector |

### The study sites

The study site was eighteen (18) Councils of the Local Government Authorities within the nine (9) regions out of 26 Regions of Tanzania Mainland from nine (9) geographical zones as summarized in Table 2 and Figure 2. The nine zones are selected to seek the country’s geographical representation. Together, these regions have a total population of 21,119,700, which represents 35.7% of the Tanzanian population. These regions are heterogeneous in population size, distribution of health facilities, human resources for health, and institutions carrying out health research activities. A similar approach has been used in major Tanzanian health studies(4,5,12); This approach provided a comprehensive understanding of the status quo for the use of health research evidence in health planning, and heterogeneous study population for random sampling hence acting as a representative snapshot of all regions in Tanzania. The study was conducted in 63 randomly selected health facilities, nine (9) Regional Referral Hospitals (RRHs), eighteen (18) District Hospitals (DHs), Eighteen (18) Health Centres (HCs), and eighteen (18) Dispensaries. To ensure the inclusion of urban and rural health facilities, stratification was conducted followed by random sampling.

**Figure 2:**
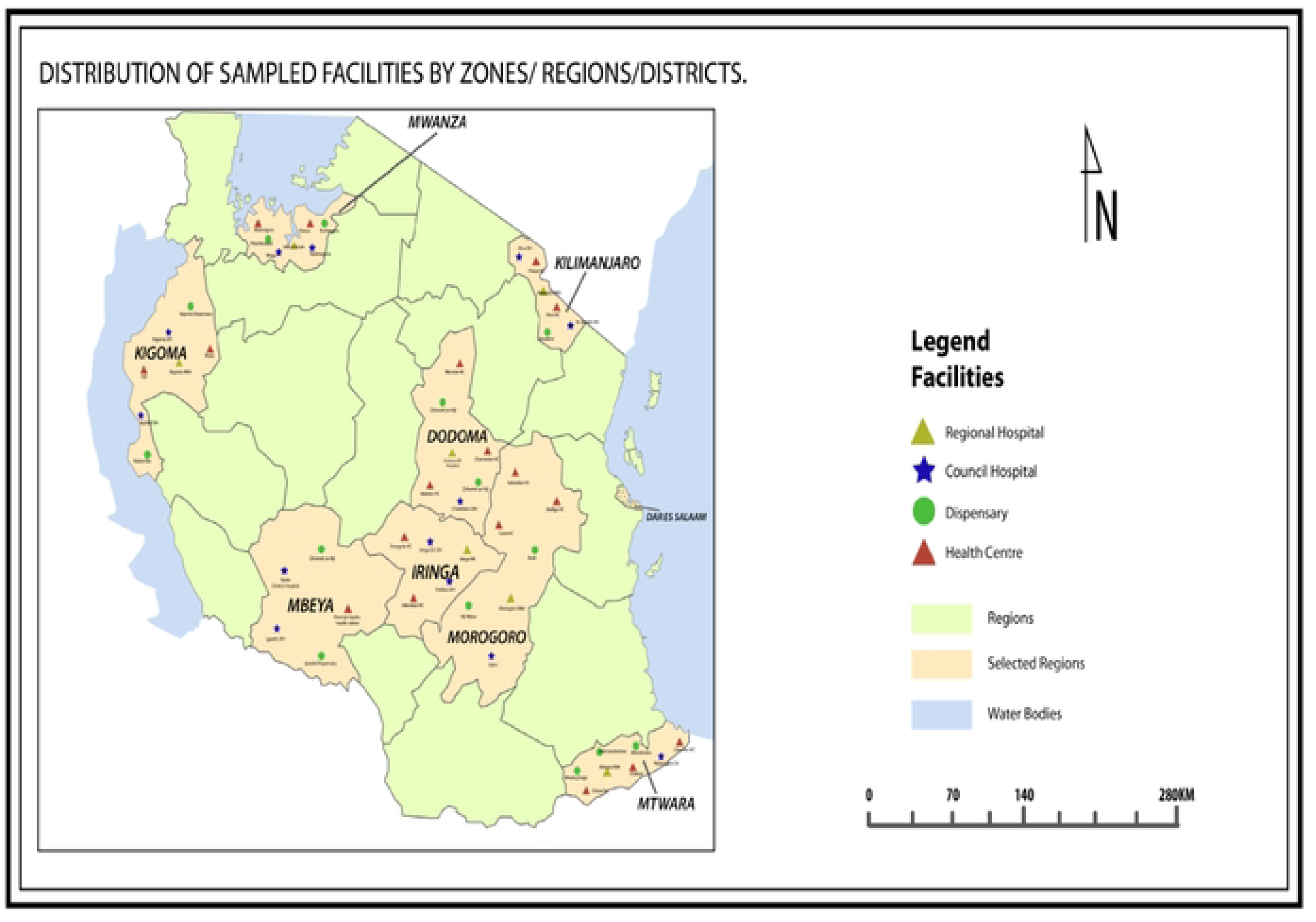
Distribution of sampled facilities by zones, regions, and districts

**Table 2:** Summary of study sites.

| ZONE | REGION | COUNCIL |
| --- | --- | --- |
| Northern |  | Siha District |
|  | Kilimanjaro | Moshi Municipal |
| Central zone |  | Chamwino District |
|  | Dodoma | Dodoma City |
| Dar es Salaam |  | Kigamboni Municipal |
|  | Dar es Salaam | Dar es Salaam City |
| Eastern |  | Morogoro Municipal |
|  | Morogoro | Gairo District |
| Western |  | Kigoma District |
|  | Kigoma | Kigoma Municipal |
| Southwest highland |  | Mbeya City |
|  | Mbeya | Kyela District |
| Lake |  | Kwimba District |
|  | Mwanza | Mwanza City |
| Southern |  | Mtwara Municipal |
|  | Mtwara | Nanyamba |
| Southern Highland | Iringa | Iringa Municipal |
|  |  | Iringa District |

### The health planning landscape in Tanzania

The health planning process in Tanzania is conducted at two levels; At the Council level, the health facility prepares its plans as well and the Council Health Management Team (CHMT) prepares its plan, whereby plans from the facility and CHMT are later consolidated to form Comprehensive Council plan (CCHP). The preparation of health facility plans is guided by the Health facility planning guidelines and CCHP guidelines.(13). At the regional level, the Regional Health Management Teams (RHMTs) prepare their plans using the RHMT planning guide and the Regional Referral Hospitals (RRHs) plans using the Comprehensive Operational Plan Guide (CHOP)(14). When the plans are completed are sent to the Ministry of Health and President’s Office-Regional Administration and Local Government for final assessment before being sent to the Ministry of Finance for funding. At all levels, we are planning to use the guidelines and the routine data collected from each level.(14).

### Study Design

The study adopts a quantitative cross-sectional design focusing on nine regions of Tanzania. (15,16).

### Sample size and Sampling procedures

The sample size involved the health planning team members from the RRHs, DHs, HCs, and Dispensaries randomly selected from the nine regions. The sample size was 422 calculated from the Cochrane formula (1977). To date, in Tanzania there is no cited reference for the percentage of the use of health research evidence in health planning therefore the marginal error of 50% was used (17).

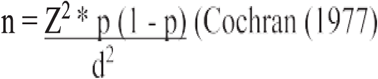

Where by

N-sample size

Z-confidence interval 95% 1.96, P-proportional from previous study d = margin of error, which is approximately 5% Thus,

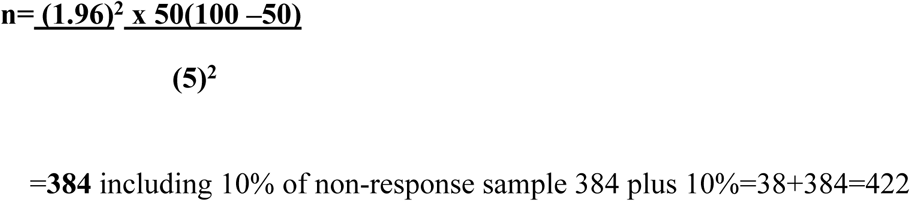

This study employed a multistage sampling technique for the selection of the study units as summarized in Figure 3. The sampling stages are zones, regions, councils, and public primary health facilities. The first stage was a random selection of one region from each of the nine Zones of the country. In the second stage, in each selected region, councils were clustered into rural and urban, and then one rural and one urban council will be selected from each region followed by a random selection of the health facilities, see Figure 3. The technique is convenient for studying large and diverse populations (5). The sampling stages were zones, regions, councils, and public primary health facilities.

**Figure 3:**
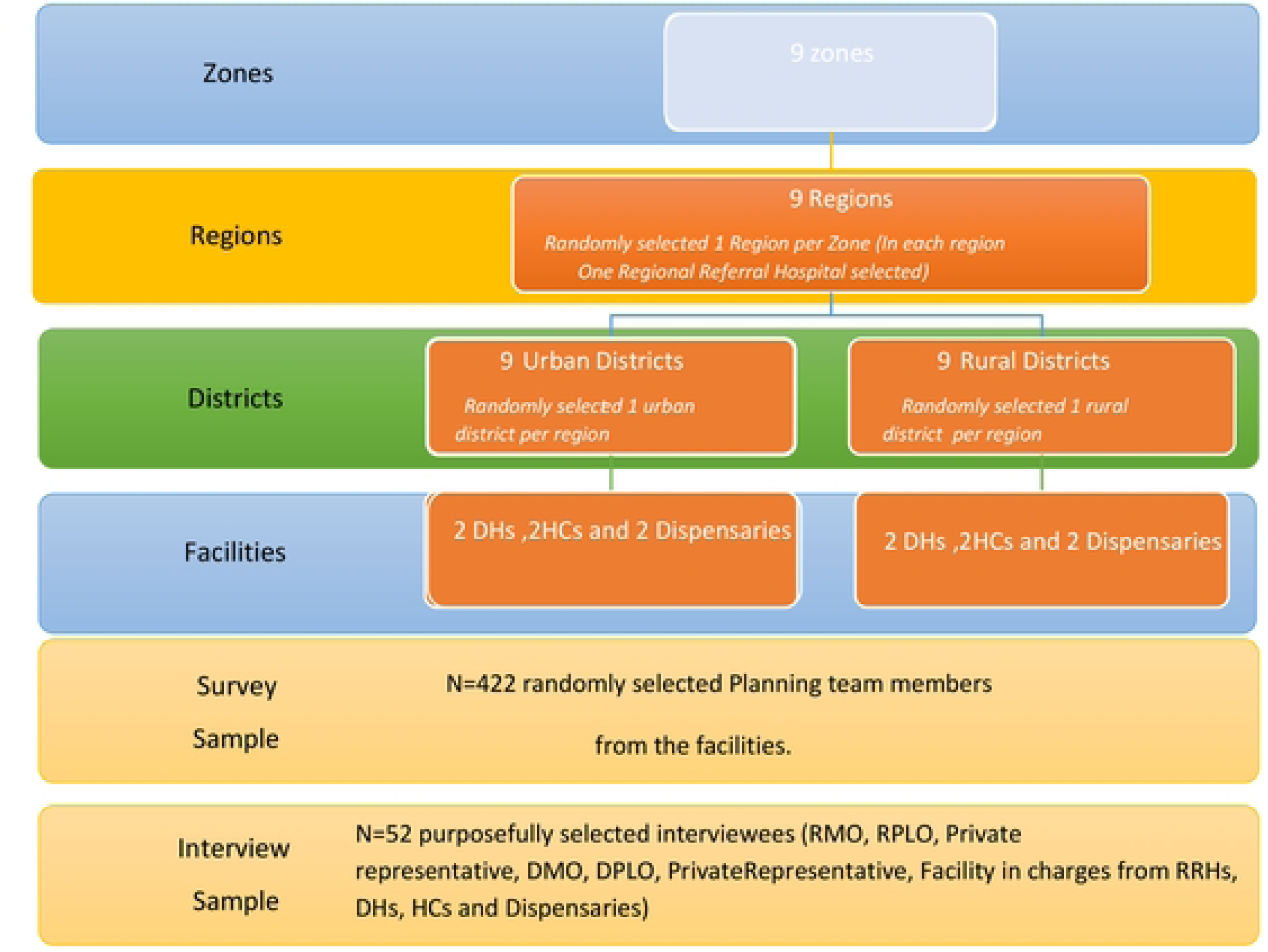

### Variables and their measurements

#### Dependent variable

The dependent variable of this study i.e. the use of health research evidence was obtained from a set of four (4) questions that were measured. The questions were a mixture of binary and multiple-choice questions. Those questions were divided into four areas. The mean score was calculated from those sets of questions; the score had a value of 0 and 1. Those respondents who scored 1 were regarded to have used the health research evidence, while those who scored 0 were regarded to have not used the health research evidence. For multiple choice questions, the respondents were provided with a set of predefined options, allowing them to select the one that best represents their response, which will capture varying degrees or categories on the use of health research evidence.

#### Independent Variables

The independent variables in this study encompass numerous factors including demographic information such as sex, age, and education level; professional attributes such as position within the health facility and years of schooling; and contextual elements including the type of health facility, stakeholders group represented district, and region. The study aimed to gain insights into the determinants that influence the use of health research evidence during health planning. The determinants had a combination of three constructs which are, capability determinants, opportunity determinants, and motivation determinants both derived from the COM-B Model.

**Capability determinants** were obtained from a set of twelve (12) Likert scale questions with a scale of 1 to 5 (1 = strongly agree, 2 = somewhat agree, 3 = neither agree nor disagree, 4 = somewhat disagree, 5 = strongly disagree) and one multiple choice question. The mean and standard deviation of the responses to assess central tendency and variability were calculated. For a multiple-choice question, the frequency distribution of each response category was analyzed. Additionally, a chi-square test was calculated to determine if the distribution of responses was significantly different from what would be expected by chance.

**Opportunity determinants,** were obtained from a set of seventeen (17) Likert scale questions with a scale of 1 to 5 (1 = strongly agree, 2 = somewhat agree, 3 = neither agree nor disagree, 4 = somewhat disagree, 5 = strongly disagree). We assigned numerical values (1 to 5) to these responses, where lower values indicate stronger agreement. Next, the mean or median was calculated of these numerical values to quantify the overall tendency. The multiple regression analysis was used to assess the relationship between the independent variable and dependent variables.

**Motivation determinants** were obtained from a set of eight (8) Likert scale questions with a scale of 1 to 5 (1 = strongly agree, 2 = somewhat agree, 3 = neither agree nor disagree, 4 = somewhat disagree, 5 = strongly disagree). We assigned numerical values (1 to 5) to these responses, where lower values indicate stronger agreement. Next, the mean or median was calculated of these numerical values to quantify the overall tendency. The multiple regression analysis was used to assess the relationship between the independent variable and dependent variables.

### Data collection procedures and tools

Quantitative data was collected face to face person by using the Swahili version of the survey after translation of the English version of the survey and document review checklist (see supplemental file) to planning team members at national, regional, council, and health facility levels. The questions from the tool were adopted and modified from previous studies (18,19). The survey collected information on the use of health research evidence and its determinants from the sampled participants using Open Data Kit software (ODK).

The document review checklist was used to guide the document review to see whether the available plans have any evidence of being prepared using health research evidence. The checklist contains a list of questions that will help the review of the plans made at the facility, council, regional, and national levels. The checklist is used to collect information on the available health plans if they are prepared by using health research evidence. Data was collected by a research assistant, who was trained for three days in data collection methods, tools, and ethics. They were selected according to their experience in the health system. Pilot testing of the tools was conducted at Dodoma RHMT and Chamwino District Council at Chamwino CHMT, Chamwino Council Hospital, Chamwino Health Centre, and Buigiri Dispensary to enhance the validity, reliability, and effectiveness of the tool in capturing the intended information.

### Data analysis

Responses were reported on all survey items in all response categories and summarized using frequencies and percentages for categorical variables and means and standard deviation for non-categorical variables were computed. Given that the outcome variable had two categories (0=Negative, 1=Positive), A binary logistic regression model was used to assess factors associated with the perception of implementers on the prime vendor system. The model results are regression parameter estimates and odds ratios (OR). The data analysis was conducted using STATA version 18, and the significance of all statistical tests was established at a 5% significance level.

### Data validity and reliability

To ensure validity and reliability, the validity of quantitative data collection tools was reviewed by independent subject matter experts from the University of Dodoma (UDOM). The reliability assessment of the questionnaire was done using the internal consistency test, with the alpha reliability coefficient being the statistic. (20,21). The range of the alpha coefficient, also known as Cronbach’s alpha was 0.71 acceptable to this study. Moreover, exploratory factor analysis to establish the construct validity of the questionnaire and pilot study conducted at Chamwino DC before field data collection to ensure clarity of the data collection tool.

### Ethics and Dissemination

The study was granted ethical approval by the University of Dodoma Ethical Committee with **Ref. No. MA.84/261/102/’A’/64/91.** Permission to conduct the study and consent to participate in the study was sought from relevant authorities and participants, respectively. Participants received information about the purpose of the study and data protection. The findings of the study were disseminated to relevant stakeholders through collaborative communication with the Ministry of Health, RHMT/CHMT, Tanzania Health Summit as well as publications to target researchers, practitioners, implementers, and policymakers.

## Results

The finding from this study is presented in three parts using tables, percentages, and charts, Part one shows the respondents’ demographic characteristics and stakeholder group. Part two shows the proportion of the respondents who reported using Health research evidence in health planning and, part three shows the association between health research use in planning and the COM-B constructs (capability, Motivation, and opportunity).

### Demographic characteristics

Table 1 describes the respondents’ characteristics and the stakeholder group they represent. There was an almost equal number of female 50.2 % and male 49.8% respondents with a mean age of 39.51 ±7.86years old. Most of them (42.7%) had undergraduate education followed by a Diploma (36.3%) and less than one-third (14.0%) had a master’s degree and above. Few (7.1%) respondents ‘had certificate-level education. Professional background results revealed that the majority 107 (25.4%) of the respondents were medical doctors followed by nurses 99 (23.5%). Physiotherapists and panning offices were very few each accounting for 0. 24% of the respondents.

### Proportion of the use of health research evidence

**Table 4** provides data on the use of health research evidence in a particular context. It shows that the majority of participants (96.7%) have used routine data, with 49.5% reporting a high extent of use. Additionally, evidence from routine data was the most commonly used type of evidence during health planning, with 98.8% of participants reporting its use. Most participants (66.2%) have used health research evidence as shown in Figure 4, and a significant majority (98.8%) have used health planning guidelines. Moreover, the majority of participants (98.8%) also indicated the importance of the use of health research evidence, with 82.6% considering it very important. These findings suggest a strong reliance on routine data and a high recognition of the importance of health research evidence in the context studied.

**Table 3:** Demographic characteristics of respondents.

| Variable | Frequency | Percent | Mean ±SD |
| --- | --- | --- | --- |
| **Age** | | | 39.51 ±7.86 |
| <25 | 3 | 0.7 |  |
| 26-35 | 159 | 37.7 |  |
| 36-45 | 156 | 37.0 |  |
| 46-55 | 93 | 22.4 |  |
| >46 | 11 | 2.6 |  |
| **Region** |  |  |  |
| Mbeya | 48 | 11.4 |  |
| Dodoma | 50 | 11.9 |  |
| Iringa | 41 | 9.7 |  |
| Mtwara | 49 | 11.6 |  |
| Mwanza | 50 | 11.85 |  |
| Kigoma | 51 | 12.1 |  |
| Kilimanjaro | 45 | 10.7 |  |
| Morogoro | 48 | 11.4 |  |
| Dar es salaam | 40 | 9.5 |  |
| **Type of health facility** |  |  |  |
| Dispensary | 37 | 8.8 |  |
| Health center | 82 | 19.4 |  |
| Council Hospital | 97 | 23.0 |  |
| Regional referral hospital | 81 | 19.2 |  |
| RHMT | 87 | 20.6 |  |
| CHMT | 38 | 9.0 |  |
| **Gender** |  |  |  |
| Male | 210 | 49.76 |  |
| Female | 212 | 50.24 |  |
| **Professional background** |  |  |  |
| Doctor | 107 | 25.36 |  |
| Nurse | 99 | 23.46 |  |
| Laboratory scientist | 36 | 8.53 |  |
| Pharmacist | 28 | 6.64 |  |
| Radiographer | 7 | 1.66 |  |
| Environmental Health officer | 13 | 3.08 |  |
| Nutrition officer | 13 | 3.08 |  |
| Social Welfare Officer | 20 | 4.74 |  |
| Physiotherapist | 1 | 0.24 |  |
| Biomedical engineer | 3 | 0.71 |  |
| Planning officer | 1 | 0.24 |  |
| Health Secretary | 38 | 9.00 |  |
| Other | 56 | 13.3 |  |
| **Stakeholders' group** |  |  |  |
| RHMT | 49 | 11.6 |  |
| HMT | 280 | 66.4 |  |
| CHMT | 90 | 21.3 |  |
| FBO | 1 | 0.2 |  |
| HGFC | 2 | 0.5 |  |
| **Highest level of education** |  |  |  |
| Certificate | 30 | 7.1 |  |
| Diploma Certificate | 153 | 36.3 |  |
| Undergraduate degree | 180 | 42.7 |  |
| Master's Degree | 56 | 13.3 |  |
| PhD | 3 | 0.7 |  |
| **The position holds currently in health planning.** |  |  |  |
| Chairperson | 30 | 7.1 |  |
| Secretary | 45 | 10.7 |  |
| Technical Advisor | 11 | 2.6 |  |
| Member | 335 | 79.4 |  |
| Other | 1 | 0.2 |  |
| **Years participate** |  |  | 5.08 ±3.96 |
| <5 | 283 | 67.1 |  |
| 6-10 | 103 | 24.4 |  |
| >10 | 36 | 8.5 |  |

**Figure 4:**
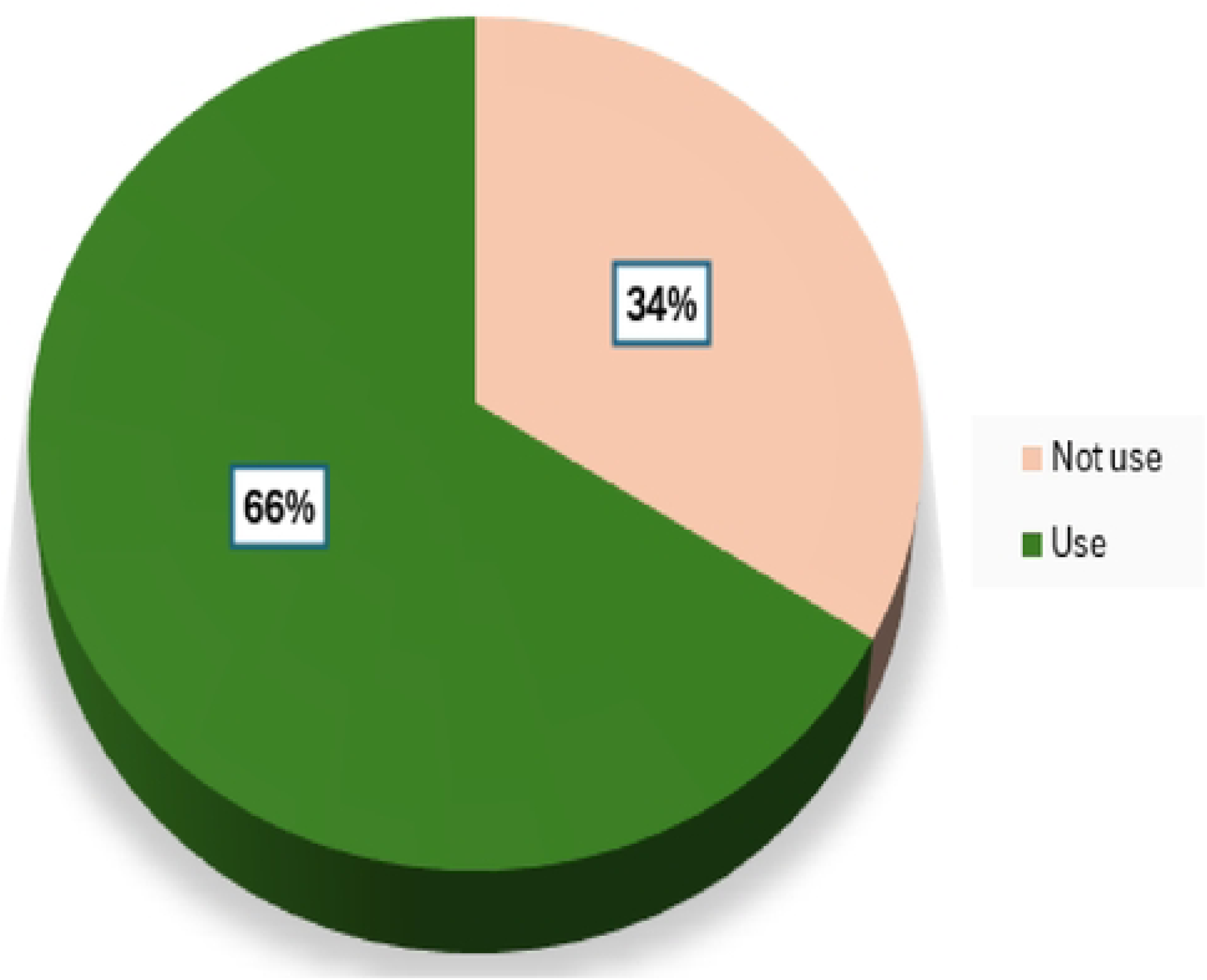
Proportion of the use of health research evidence.

**Table 4:** The proportion of the use of health research evidence during health planning.

| Variable | Frequency | Percent |
| --- | --- | --- |
| **Ever used routine data** |  |  |
| No | 14 | 3.3 |
| Yes | 408 | 96.7 |
| **Extent of use of routine data** |  |  |
| Low | 41 | 10.1 |
| Medium | 165 | 40.4 |
| High | 202 | 49.5 |
| **Type of evidence used during health planning (Multiple response)** |  |  |
| Evidence from routine data | 403 | 98.8 |
| Policy documents | 280 | 68.6 |
| Research publications (general) | 133 | 32.6 |
| Systematic reviews/ meta-analysis | 41 | 10.1 |
| Randomized Control trials | 17 | 4.2 |
| Experimental studies | 30 | 7.4 |
| Non-experimental studies | 20 | 4.9 |
| Expert opinions | 271 | 66.4 |
| Policy beliefs | 162 | 39.7 |
| Other | 17 | 4.2 |
| **Use of health research evidence** |  |  |
| No | 138 | 33.8 |
| Yes | 270 | 66.2 |
| **Use of the Health planning guidelines** |  |  |
| No | 5 | 1.2 |
| Yes | 403 | 98.8 |
| **Use of the Ruling party manifesto** |  |  |
| No | 44 | 10.8 |
| Yes | 364 | 89.2 |
| **Use of the Policy documents** |  |  |
| No | 137 | 33.6 |
| Yes | 271 | 66.4 |
| **Importance of the use of health research evidence** |  |  |
| No | 5 | 1.2 |
| Yes | 403 | 98.8 |
| **Level of importance of the use of health research evidence** |  |  |
| Very unimportant | 5 | 1.2 |
| Unimportant | 2 | 0.5 |
| Neutral | 14 | 3.4 |
| Important | 50 | 12.3 |
| Very important | 337 | 82.6 |

### The factors associated with the use of health research evidence during health planning

The table presents the results of a binary logistic regression analysis examining factors associated with the use of research evidence during health planning. The analysis includes variables such as age, region, type of health facility, gender, professional background, stakeholder group, highest level of education, position held in health planning, and years of participation. The adjusted odds ratios (OR), confidence intervals (CI), and p-values are provided to indicate the strength and statistical significance of associations.

#### Significant Variables and Interpretation

- **Region**: The analysis compares various regions to the reference category, Kilimanjaro. The odds ratios (OR) show how likely individuals from other regions are to use health research evidence in health planning compared to those from Kilimanjaro. For instance, if a region has an OR greater than 1, individuals from that region are more likely to use research evidence than those from Kilimanjaro. If the OR is less than 1, they are less likely. The p-values indicate whether these differences are statistically significant.
- **Level of Education**: Compared to individuals with a diploma (reference category), those with higher education levels were more likely to use research evidence:

- Undergraduate degree (OR = 2.396, CI: [1.094, 5.249], p-value = 0.0290): Those with an undergraduate degree are over twice as likely to use research evidence compared to diploma holders.
- Master’s degree (OR = 2.278, CI: [0.921, 5.631], p-value = 0.0747): Similar to undergraduates, master’s degree holders are also more likely to use research evidence in health planning although were not significant.

These OR values reflect the increased likelihood of individuals with higher education levels using research evidence compared to the reference group, with p-values indicating the strength of this relationship.

- Years of Participation:

- 6–10 years (OR = 1.092, CI: [0.677,1.761], p-value = 0.7184): Individuals with 6–10 years of participation in health planning show a higher likelihood of using research evidence compared to those with fewer years, though this was not statistically significant.
- More than 10 years (OR = 1.399, CI: [0.648,3.019], p-value = 0.3924): Those with over 10 years of participation have a 40% higher likelihood of using research evidence compared to those with fewer years of experience, and this difference was also not statistically significant.

The adjusted odds ratios reflect the likelihood of each category of the variable influencing the use of research evidence, controlling for other variables in the model. Significance is determined by both the size of the OR and the p-value, with confidence intervals providing a range within which the true effect likely falls.

#### Capability: The knowledge and skills of health research evidence use

Table 6 presents the findings from a survey on the capacity to use health research evidence (knowledge and skills). Each row in the table represents a different variable related to the use of health research evidence, and the columns display the responses categorized into five levels of importance: Very Unimportant (VUIM), Unimportant (UIM), Neutral, Important (IM), and Very Important (VIM).

**Table 5:** Binary logistic regression for factors associated with the use of research evidence during health planning.

| Variable | Not use | Use | Unadjusted logistic analysis |  | Adjusted logistic analysis |  |
| --- | --- | --- | --- | --- | --- | --- |
|  | N (%) | N (%) | OR [95%CI] | p-value | OR [95%CI] | p-value |
| **Age** |  |  |  |  |  |  |
| <35 | 52(32.1) | 110(67.9) | ref |  |  |  |
| 36-45 | 56(35.9) | 100(64.1) | 0.844[0.530,1.343] | 0.4747 |  |  |
| >45 | 35(33.7) | 69(66.4) | 0.932[0.552,1.573] | 0.7919 |  |  |
| **Region** |  |  |  |  |  |  |
| Kilimanjaro | 26(57.8) | 19(42.2) | ref |  | ref |  |
| Mbeya | 17(35.4) | 31(64.6) | 2.495[1.081,5.760] | 0.0322 | 2.604[1.074,6.315] | 0.0342 |
| Dodoma | 17(34.0) | 33(66.0) | 2.656[1.156,6.104] | 0.0214 | 2.514[1.053,6.001] | 0.0378 |
| Iringa | 10(24.4) | 31(75.6) | 4.242[1.680,10.71] | 0.0022 | 4.707[1.772,12.499] | 0.0019 |
| Mtwara | 21(42.9) | 28(57.1) | 1.825[0.805,4.137] | 0.1499 | 1.801[0.727,4.466] | 0.2039 |
| Mwanza | 12(24.0) | 38(76.0) | 4.333[1.801,10.43] | 0.0011 | 4.794[1.893,12.140] | 0.0009 |
| Kigoma | 10(19.6) | 41(80.4) | 5.611[2.259,13.94] | 0.0002 | 5.922[2.224,15.769] | 0.0004 |
| Morogoro | 17(35.4) | 31(64.6) | 2.495[1.081,5.760] | 0.0322 | 2.180[0.901,5.274] | 0.0839 |
| Dar es salaam | 13(32.5) | 27(67.5) | 2.842[1.170,6.904] | 0.0211 | 3.229[1.211,8.611] | 0.0192 |
| **Type of health facility** |  |  |  |  |  |  |
| Dispensary | 17(46.0) | 20(54.1) | ref |  | ref |  |
| Health center | 25(30.5) | 57(69.5) | 1.938[0.871,4.311] | 0.1048 | 2.029[0.829,4.964] | 0.1211 |
| Council Hospital | 45(46.4) | 52(53.6) | 0.982[0.459,2.100] | 0.9631 | 0.809[0.323,2.026] | 0.6511 |
| RRH | 19(23.5) | 62(76.5) | 2.774[1.214,6.335] | 0.0155 | 2.663[1.895,7.920] | 0.0383 |
| RHMT | 27(31.0) | 60(69.0) | 1.889[0.857,4.163] | 0.1147 | 0.350[0.031,3.902] | 0.3932 |
| CHMT | 10(26.3) | 28(73.7) | 2.380[0.903,6.273] | 0.0795 | 5.647[0.602,52.94] | 0.3167 |
| **Gender** |  |  |  |  |  |  |
| Male | 71(33.8) | 139(66.2) | 1.007[0.673,1.507] | 0.9736 |  |  |
| Female | 72(34.0) | 140(66.0) | ref |  |  |  |
| **Professional background** |  |  |  |  |  |  |
| Doctor | 37(34.6) | 70(65.4) | ref |  |  |  |
| Nurse | 35(35.4) | 64(64.7) | 0.967[0.545,1.714] | 0.9073 |  |  |
| Lab scientist & Pharmacist | 25(35.2) | 46(64.8) | 0.973[0.518,1.825] | 0.9310 |  |  |
| Non-medical | 28(31.5) | 61(68.5) | 1.152[0.633,2.096] | 0.6444 |  |  |
| Other | 18(32.1) | 38(67.7) | 1.116[0.561,2.220] | 0.7548 |  |  |
| **Stakeholders' group** |  |  |  |  |  |  |
| RHMT | 13(26.5) | 36(73.5) | 1.609[0.816,3.171] | 0.1697 | 0.843[0.177,4.012] | 0.8297 |
| CHMT | 26(28.9) | 64(71.1) | 1.430[0.854,2.396] | 0.1741 | 5.647[0.602,52.93] | 0.1295 |
| HMT | 104(36.8) | 179(63.3) | ref |  | ref |  |
| **Highest level of education** |  |  |  |  |  |  |
| Certificate | 15(50.0) | 15(50.0) | ref |  | ref |  |
| Diploma | 57(37.3) | 96(62.8) | 1.684[0.767,3.700] | 0.1943 | 1.609[0.670,3.867] | 0.2874 |
| Undergraduate | 53(29.4) | 127(70.6) | 2.396[1.094,5.249] | 0.0290 | 1.824[0.710,4.685] | 0.2120 |
| Master's Degree | 18(30.5) | 41(69.5) | 2.278[0.921,5.631] | 0.0747 | 2.082[0.684,6.337] | 0.1968 |
| **Position holds currently in health planning** |  |  |  |  |  |  |
| Secretary | 19(42.2) | 26(57.8) | ref |  | ref |  |
| Chairperson | 8(26.7) | 22(73.3) | 2.009[0.738,5.475] | 0.1724 | 1.933[0.647,5.780] | 0.2381 |
| Technical | 4(36.4) | 7(63.6) | 1.279[0.327,5.000] | 0.7236 | 0.789[0.173,3.599] | 0.7600 |
| Member | 112(33.3) | 224(66.7) | 1.462[0.776,2.754] | 0.2404 | 1.328[0.651,2.708] | 0.4354 |
| **Years participate** |  |  |  |  |  |  |
| <5 | 99(35.0) | 184(65.0) | ref |  |  |  |
| 6-10 | 34(33.0) | 69(67.0) | 1.092[0.677,1.761] | 0.7184 |  |  |
| >10 | 10(27.8) | 26(72.2) | 1.399[0.648,3.019] | 0.3924 |  |  |

**Table 6:**
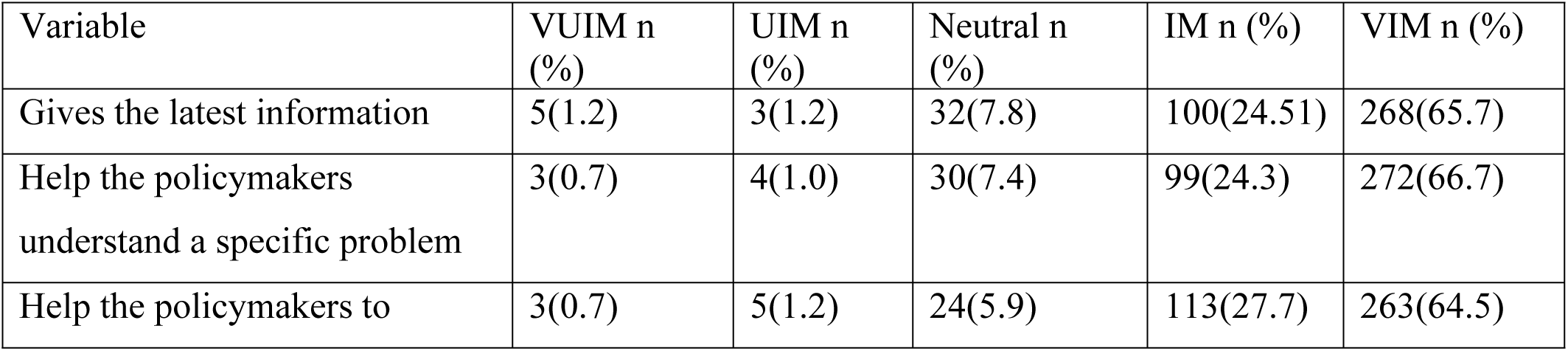
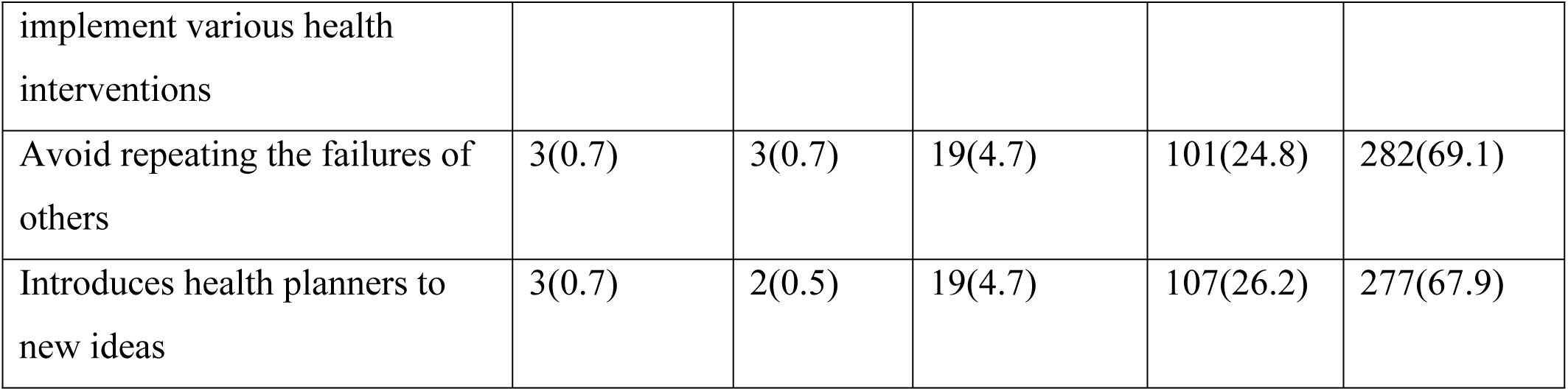
Knowledge and skills of health research evidence use.

From the data, it’s clear that the majority of respondents perceive the use of health research evidence as very important or important across the various variables. In the variable “Gives the latest information,” the majority of respondents rated it as Very Important (65.7%) or Important (24.5%). Similarly, in the variable “Help the policymakers understand a specific problem,” a substantial proportion of respondents indicated that it is Very Important (66.7%) or Important (24.3%). Overall, the data suggests that there is a strong recognition of the significance of health research evidence in various aspects of policymaking and intervention implementation.

Table 7 provides information on the obstacles hindering the utilization of health research evidence. The variables include Lack of necessary knowledge and training in research, Inadequate human and non-human resources, Difficulty in accessing health research evidence, Perceptions that health research is for academic purposes, Lack of involvement in research activities, and Traditional ways of planning, as well as the minimal usage of health research evidence among planning team members at the regional and council levels, and lack of dissemination.

**Table 7:**
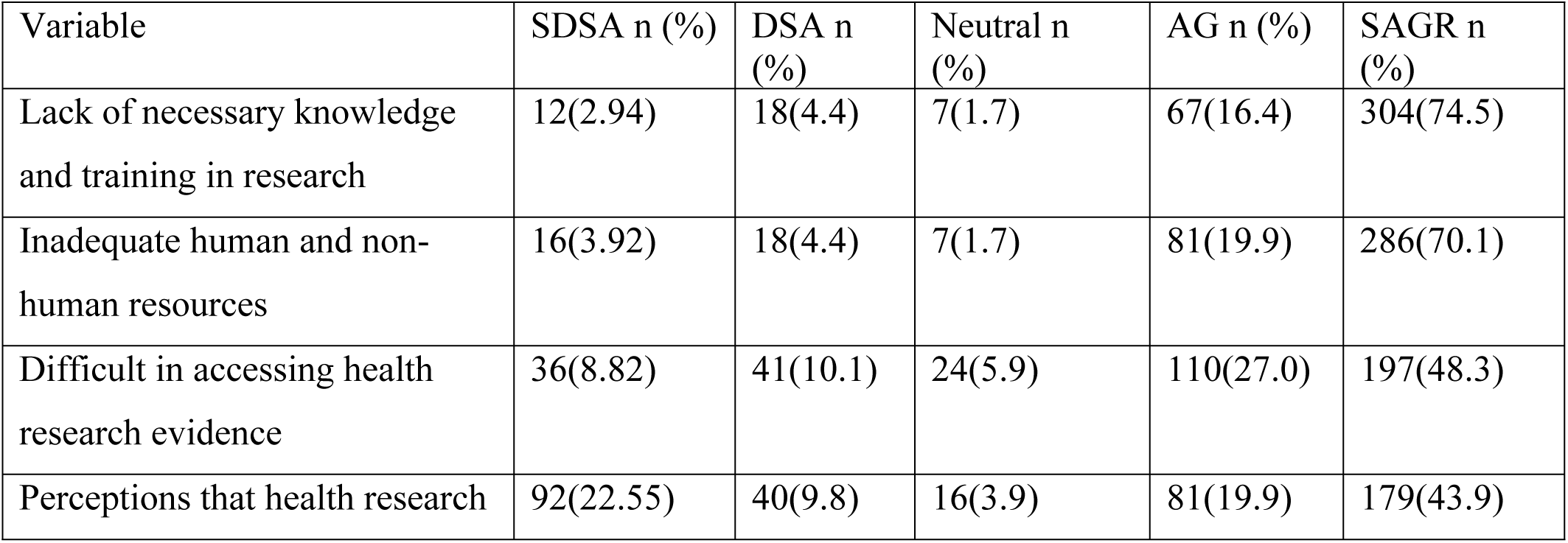
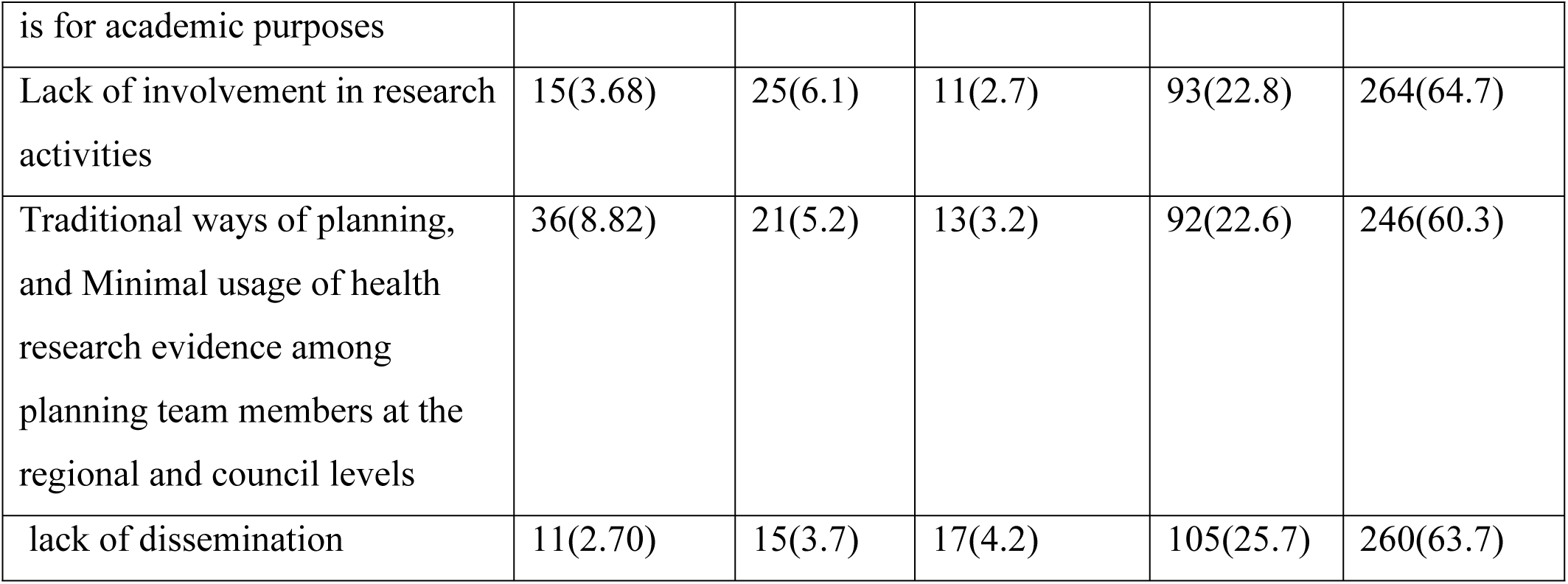
Obstacles that hinder the utilization of health research evidence.

The data is presented in terms of the number and percentage of participants who strongly disagree (SDSA), disagree (DSA), are neutral, agree (AG), and strongly agree (SAGR) with each obstacle. This data provides insight into the specific challenges that impact the utilization of health research evidence within the context of the study. The highest percentage is for “Perceptions that health research is for academic purposes” at 22.6%, and the lowest percentage is for “Lack of necessary knowledge and training in research” at 2.9%.

### Opportunities for the use of health research evidence

Table 8 indicates the opportunities for the use of health research evidence, helping stakeholders to identify areas for improvement and investment to optimize the use of health research evidence. The table presents the opportunities for utilizing health research evidence, categorized into physical and social opportunities.

**Table 8:** Opportunities for the use of health research evidence.

| Variable | SDSA n (%) | DSA n (%) | Neutral n (%) | AG n (%) | SAGR n (%) |
| --- | --- | --- | --- | --- | --- |
| **Physical opportunities** |  |  |  |  |  |
| Enabling the utilization of information generated from the healthcare system | 6(1.4) | 9(2.1) | 18(4.3) | 130(30.8) | 259(61.4) |
| Availability of research coordinators | 9(2.1) | 18(4.3) | 22(5.2) | 93(22.0) | 280(66.4) |
| Availability of equipment | 4(1.0) | 16(3.8) | 21(5.0) | 88(20.9) | 293(69.4) |
| Availability of internet | 3(0.7) | 9(2.1) | 19(4.5) | 69(16.4) | 322(76.3) |
| Availability of planned budget for research | 11(2.6) | 17(4.0) | 27(6.4) | 64(15.2) | 303(71.8) |
| Presence of symposiums for research discussions | 9(2.1) | 17(4.0) | 23(5.5) | 106(25.1) | 267(63.3) |
| **Social opportunities** |  |  |  |  |  |
| The presence of disparities among different populations allows for more equitable | 10(2.4) | 24(5.7) | 38(9.0) | 147(34.8) | 203(48.1) |
| The presence of community members in the health planning teams | 11(2.6) | 28(6.6) | 55(13.0) | 119(28.2) | 209(49.5) |
| Improved health literacy in the society, empowering individuals to make informed | 4(1.0) | 19(4.5) | 18(4.3) | 145(34.4) | 236(55.9) |

For physical opportunities, the table highlights the availability of resources such as research coordinators, equipment, internet access, a planned budget for research, and the presence of symposiums for research discussions. The percentages indicate the distribution of responses, ranging from strongly disagree to strongly agree, reflecting the varying levels of agreement among the respondents regarding these physical opportunities. In the physical opportunities category, the highest percentage is 76.3%, representing the availability of the internet, while the lowest percentage is 0.7%, indicating the availability of Internet.

In the social opportunities section, the table emphasizes the presence of disparities among different populations, the involvement of community members in health planning teams, and the enhancement of health literacy in society. The percentages provide an insight into the attitudes and perceptions of the respondents toward these social opportunities. In the social opportunities category, the highest percentage is 55.9%, reflecting improved health literacy in society, and the lowest percentage is 2.4%, which represents the presence of disparities among different populations allowing for more equitable representation.

### Motivations for the use of health research evidence

Table 9, based on the data presented, we can observe that the automatic motivations for the use of health research evidence are primarily centered around the availability of transparency and accountability mechanisms 66.4% strongly agreeing, and 23.2% agreeing with the statement availability of job training 73.2% strongly agreeing and 21.3% agreeing respectively. Provision of incentives to health planners, with 72.4% strongly agreeing and 15.4% agreeing respectively. In contrast, reflective motivations such as continuous quality improvement in the healthcare system and the presence of active stakeholder engagement in health planning also garnered significant support, with 63.51% and 59.48% strongly agreeing or agreeing, respectively. This indicates a strong emphasis on the proactive involvement of various stakeholders and the continuous enhancement of the healthcare system based on research evidence.

**Table 9:** Motivations of the use of health research evidence.

| Variable | SDSA n (%) | DSA n (%) | Neutral n (%) | AG n (%) | SAGR n (%) |
| --- | --- | --- | --- | --- | --- |
| **Automatic motivations** |  |  |  |  |  |
| Provision of incentives to health planners | 19(4.5) | 21(5.0) | 13(3.1) | 65(15.4) | 304(72.0) |
| Availability of job training | 3(0.7) | 4(1.0) | 16(3.8) | 90(21.3) | 309(73.2) |
| Availability of Short-term courses | 5(1.2) | 13(3.1) | 21(5.0) | 75(17.8) | 308(73.0) |
| Availability of Long-term course | 18(4.3) | 24(5.7) | 24(5.7) | 105(24.9) | 251(59.5) |
| **Reflective motivations** |  |  |  |  |  |
| Presence of interaction between policymakers, implementers, researchers, and academicians | 3(0.7) | 17(4.0) | 21(5.0) | 111(26.3) | 270(64.0) |
| Presence of active stakeholder engagement in health planning | 6(1.4) | 16(3.8) | 33(7.8) | 116(27.5) | 251(59.5) |
| Continuous quality improvement in the healthcare system | 1(0.2) | 10(2.4) | 12(2.8) | 131(31.0) | 268(63.5) |
| Availability of transparency and accountability mechanisms | 5(1.2) | 12(2.8) | 27(6.4) | 98(23.2) | 280(66.4) |

### Findings from document review checklist

The analyzed data from Table 10 indicates that the majority of planning teams incorporate research evidence into their health plans. A substantial 97.8% of respondents confirmed having a current fiscal year health plan, and 98.9% reported having a planning guideline. Most plans include key health frameworks and guides, with 70.8% having the CCHP, 83.2% using the HF Planning Guide, and 89.9% incorporating situational analysis. Scientific engagement is evidenced by 75.3% of teams receiving invitation letters for conferences. However, challenges remain, as only 22.5% have a budget for research, and only 23.6% allocate budget specifically for research within the health plan. Additionally, just 28.1% include a research coordinator in the planning team, and 43.8% of the plans are well-cited. This highlights a strong foundation in research usage but also gaps in financial and personnel support for research-driven planning.

**Table 10:**
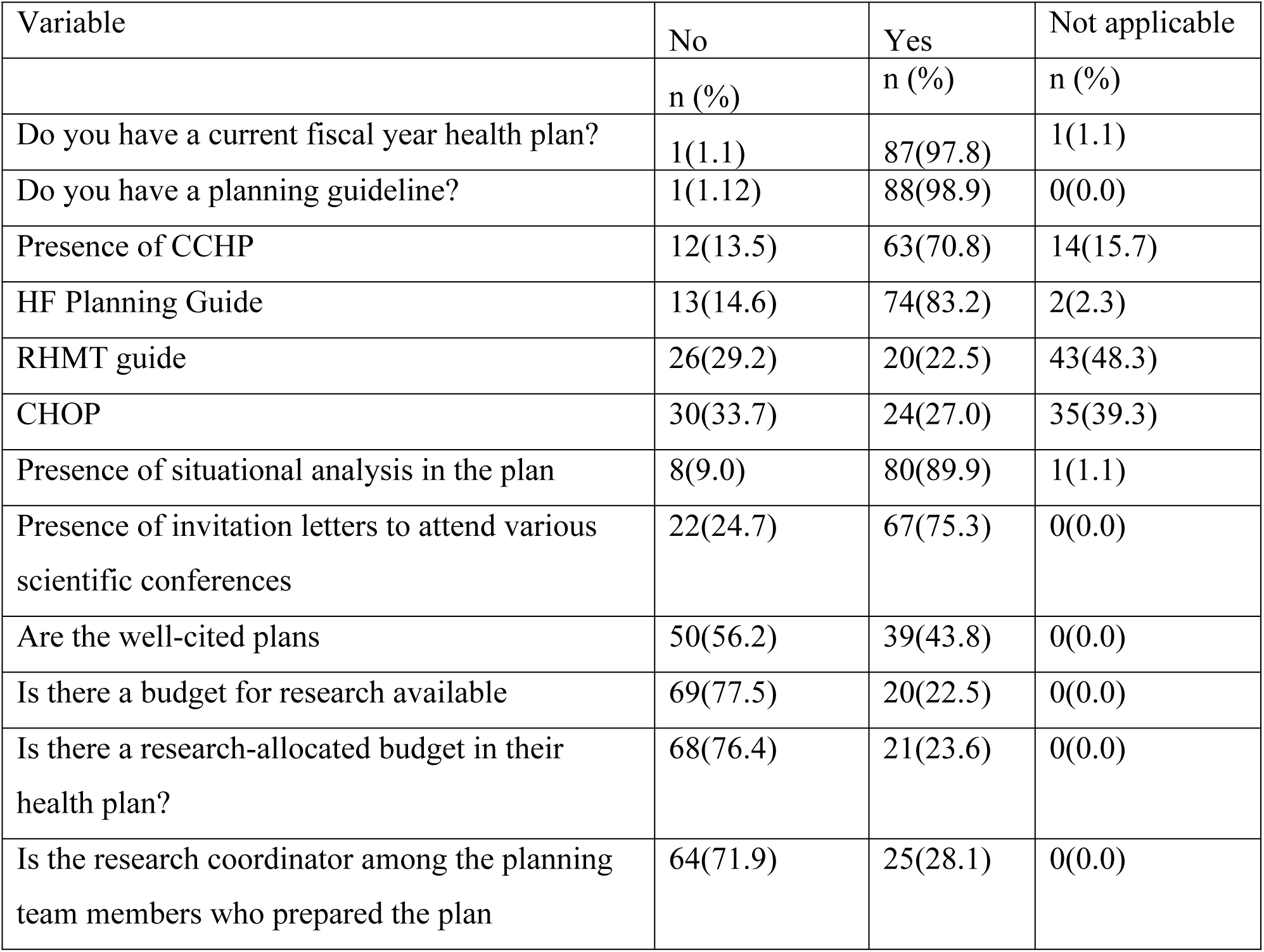
looking for evidence that planning teams use health research evidence in their plans.

| Variable | No | Yes | Not applicable |
| --- | --- | --- | --- |
|  | n (%) | n (%) | n (%) |
| Do you have a current fiscal year health plan? | 1(1.1) | 87(97.8) | 1(1.1) |
| Do you have a planning guideline? | 1(1.12) | 88(98.9) | 0(0.0) |
| Presence of CCHP | 12(13.5) | 63(70.8) | 14(15.7) |
| HF Planning Guide | 13(14.6) | 74(83.2) | 2(2.3) |
| RHMT guide | 26(29.2) | 20(22.5) | 43(48.3) |
| CHOP | 30(33.7) | 24(27.0) | 35(39.3) |
| Presence of situational analysis in the plan | 8(9.0) | 80(89.9) | 1(1.1) |
| Presence of invitation letters to attend various scientific conferences | 22(24.7) | 67(75.3) | 0(0.0) |
| Are the well-cited plans | 50(56.2) | 39(43.8) | 0(0.0) |
| Is there a budget for research available | 69(77.5) | 20(22.5) | 0(0.0) |
| Is there a research-allocated budget in their health plan? | 68(76.4) | 21(23.6) | 0(0.0) |
| Is the research coordinator among the planning team members who prepared the plan | 64(71.9) | 25(28.1) | 0(0.0) |

## Discussion

This study represents the first comprehensive analysis of the use of health research evidence in Tanzania. It aimed to analyze the determinants of the use of health research evidence by deep-diving into the three determinants influencing behavior i.e. Capability, Opportunity, and Motivation. The findings from this study indicated that all three determinants significantly impact the ability of health planners to use health research evidence during health planning. However, we found that the use of research evidence is associated with having a degree, more than 10 years of involvement in health planning activities, and prior knowledge in research. We found several facilitator determinants such as organizational support infrastructures like, the availability of computers and the internet, the availability of research experts and research coordinators, availability of academic institutions that give a chance for dissemination of research. On the other hand, barrier determinants included lack of human resources, lack of funding for research activities, lack of necessary knowledge and training, perceptions that research is for academic purposes, and lack of dissemination.

### Proportion of the use of health research evidence

The findings of this study on the proportion of the use of health research evidence demonstrate a high level of reliance on health research evidence, particularly routine data, during health planning. The majority of participants (96.7%) reported using routine data, with nearly half (49.5%) using it extensively, which is consistent with findings from similar studies. For instance, a study by Lavis et al (22) found that routine data and health statistics are frequently utilized in policy-making and health-planning processes, underscoring the central role of such data in evidence-based decision-making. Moreover, the widespread use of health planning guidelines (98.8%) and the high proportion of participants (98.8%) recognizing the importance of health research evidence align with findings from studies on the use of research evidence in health decision-making process which emphasize the role of guidelines and evidence in shaping health policies (23,24). The high rate of health research evidence use (66.2%) in this study also mirrors the findings which reported similar levels of research evidence use in healthcare settings(25). These results highlight the growing recognition and integration of research evidence in health planning, reflecting broader trends in evidence-informed policymaking across health systems in Tanzania and globally.

### Capability: The knowledge and skills of health research evidence use

Explicitly, capability determinants such as knowledge and skills were found to be critical, with health planning team members who had higher levels of education (Undergraduate, a master level, and above) and access to training more likely to use health research evidence to almost 70%, with the capability constraints faces more those with certificate and diploma level, lack of necessary knowledge and training in research accounts up to 70%, being the most barrier to utilization of health research evidence, inadequate human and non-human resources, difficult in accessing research evidence, perception that research is for academic purposes and lack of dissemination. These findings are consistent with systematic reviews conducted in other low- and middle-income countries, where similar capability constraints such as poor access to good quality relevant research, and lack of timely research output have been identified as major barriers to evidence-based health planning (26,27).

### Opportunities for the use of health research evidence

In comparing our results with existing literature on Opportunity determinants (both physical and social), it is evident that the availability of resources and institutional support plays a crucial role in enabling the use of research evidence. Our study revealed that health planning teams with better access to physical opportunities such as health information system, availability of research coordinators, equipment, internet, research budgets, presence of symposiums for research discussion, the existence of academic institutions such as universities, and social opportunities like the presence of community members representatives within the health planning teams and improved health literacy in the community were more inclined to utilize research findings in their work. This aligns with findings from two studies conducted in LMICs (28,29), which highlighted the importance of institutional support in facilitating the use of research evidence in health policy. However, our study also noted a distinct challenge in the Tanzanian context with limited access to relevant health research evidence due to infrastructural constraints, a barrier less emphasized in studies from more resource-rich settings (30,31).

### Motivations for the use of health research evidence

Particularly the automatic motivations to utilize health research evidence were also found to be significant, especially on the provision of incentives to health planners to greater than 72%.

Other motivations include job training, and short and long-term courses that influenced the motivations to use the health research evidence. The automatic motivation factors included the presence of interaction between policymakers, implementers, researchers, and academicians, and continuous quality improvement agenda in the health sector. Health planners who perceived the use of research evidence as beneficial and aligned with their professional goals were more likely to engage with it actively. This is in line with the COM-B model, which underscores the role of motivation in behavior change (10). Our findings add to the body of literature by highlighting the specific motivational barriers in Tanzania, such as a lack of a clear roadmap or framework for the utilization of health research evidence, which can reduce the enthusiasm for utilizing research in health planning (32)(33).

### Document review checklist

The findings from the documents review checklist revealed a high level of integration of research evidence in health planning aligning with similar studies that underscore the importance of research-informed frameworks in public health planning (34)(35). The near-universal adoption of fiscal-year health plans (97.8%) and planning guidelines (98.9%) is consistent with global trends in evidence-based health planning, which emphasizes standardized planning processes (36). The presence of documents like CCHP and HF Planning guide mirrors findings from an urban strategic planning using blue-green infrastructure (37), who highlighted their role in improving health outcomes, the limited financial resources dedicated to health research in this study 22.5% suggests a common barrier. Similarly, the low inclusion of research coordinators (28.1%) echoes observations by a study designed for research approach during learning (38) on the limited interdisciplinary engagement in health planning. These gaps in budget allocation and personnel highlight persistent structural challenges to fully operationalizing research evidence-backed plans, despite a broad commitment to evidence-based frameworks.

The study provided valuable insights into the factors influencing the use of health research evidence during health planning in Tanzania. The results from the binary logistic regression analysis highlight significant associations between region, level of education, and years of participation in health planning. Notably, region was a significant predictor, with participants from regions other than Kilimanjaro demonstrating varying likelihoods of using research evidence, as indicated by the odds ratios. Educational attainment also emerged as a key determinant; individuals with undergraduate degrees were over twice as likely to use health research evidence compared to those with a diploma, with statistically significant results (p-value = 0.029). However, while master’s degree holders showed a similar trend, the relationship was not statistically significant (p-value = 0.0747). The analysis of years of participation revealed that those with more than 10 years of experience were 40% more likely to use health research evidence, though this association was not statistically significant. These findings align with other studies that emphasize the importance of education and experience in evidence-based health planning (24,26). The adjusted odds ratios and confidence intervals provide a nuanced understanding of the strength and variability of these associations, contributing to a growing body of literature on the determinants of research utilization in public health policy and planning.

The implications of these findings are essential for improving health planning in Tanzania. To enhance the use of health research evidence, interventions must address all three determinants: enhancing the capability of health planners through capacity building, improving opportunities by ensuring better access to human and non-human resources and organizational support, and fostering motivation by recognizing and rewarding health planners that use evidence-based decision-making. These strategies align with recommendations from similar studies in other LMICs, suggesting that a multifaceted approach including the development of middle range theory (MRT) outlining the main facilitators for utilization of evidence for malaria treatment policy in Uganda and the importance of measuring total brain activity in neuroimaging are essential for promoting the use of research in health planning (39)(40).

## Conclusion

This study provides a comprehensive assessment of the determinants influencing the use of health research evidence in Tanzania, focusing on capability, opportunity, and motivation. The findings demonstrate that all three determinants significantly impact the behavior of health planners, highlighting those factors such as higher education levels, years of experience in health planning, and prior research knowledge contribute to greater use of health research evidence. Organizational infrastructure and institutional support further facilitate this process, while barriers include insufficient human resources, lack of funding, inadequate training, and perceptions that research is solely for academic purposes.

The study also reveals a high reliance on routine data and health planning guidelines, indicating the growing recognition of research evidence in health decision-making. However, addressing the barriers to research evidence use will require targeted interventions that strengthen the capability of health planners, improve access to resources and institutional support, and enhance motivation through incentives and professional development.

Future research should explore the long-term impact of these interventions, as well as contextual factors like political will and leadership, to fully understand how these determinants collectively shape health planning behavior. A multifaceted approach i.e. a generic framework for strengthening Knowledge Management in Tanzania (KMT) that will have a triad pathway: Capacity building on healthcare workers and researchers, infrastructure development, and funding that will improve access to health research database and resources within the healthcare system, and organizational support through the establishment of guidelines for the use of health research evidence in the decision-making process. This is essential for promoting the use of health research evidence, ultimately improving health outcomes in Tanzania and other low- and middle-income countries with similar challenges (41,42)

## Abbreviations

CCHP: Comprehensive Council Health Planning

CHOP: Comprehensive Hospital Operational Plan

LGAs: Local Government Authorities

LMICs: Lower-and Middle-Income Countries

MoH: Ministry of Health

PO-RALG: President’s Office-Regional Administration and Local Government

RHMT: Regional Health Management Team

SDGs: Sustainable Development Goals

UHC: Universal Health Coverage

WHO: World Health Organization

## Data Availability

All data produced in the present study are available upon reasonable request to the authors

## Acknowledgments

We would like to acknowledge the contribution of the University of Dodoma faculty members from the School of Nursing and Public Health for their input in the process of development of this manuscript. We would like to express our sincere thanks to the study facilities, the RHMTs, and the CHMTs for their support. We are grateful to the research assistants (Edmund Bunyaga, Theresia Ngungulu, Jestina Nyondo, Devina Mafuta Micky Masanyaji, and Ally Kinyaga) for their assistance during data collection.

## Author Contributions

PK; Conceptualization, design of the work, data collection and analysis, preparing the manuscript draft, developing the methodology, results, discussion, and conclusion. RM, Conceptualization, critically reviewing the methodology and critically reviewing the manuscript, AK, Conceptualization, critically reviewing the methodology. All authors revised the work and approved the paper to be submitted for publication.

## REFERENCES

1. Khalid FJI. The Impact of Poor Planning and Management on the Duration of Construction Projects. Multi-Knowl Electron Compr J Educ Sci Publ [Internet]. 2019;(2):1–20. Available from: www.mecsj.com

2. Onwujekwe O, Etiaba E, Mbachu C, Arize I, Nwankwor C, Ezenwaka U, et al. Does improving the skills of researchers and decision-makers in health policy and systems research lead to enhanced evidence-based decision making in Nigeria?—A short term evaluation. Oelke N, editor. PLOS ONE [Internet]. 2020 Sep 3 [cited 2024 Feb 16];15(9):e0238365. Available from: https://dx.plos.org/10.1371/journal.pone.0238365

3. Henriksson DK, Peterson SS, Waiswa P, Fredriksson M. Decision-making in district health planning in Uganda: Does use of district-specific evidence matter? Health Res Policy Syst. 2019 Jun 6;17(1).

4. Kagoma P, Mongi R, Kapologwe NA, Kengia J, Kalolo A. Health research evidence: its current usage in health planning, determinants and readiness to use knowledge translation tools among health planning teams in Tanzania—an exploratory mixed-methods study protocol. BMJ Open [Internet]. 2024 Jun [cited 2024 Jun 27];14(6):e081517. Available from: https://bmjopen.bmj.com/lookup/doi/10.1136/bmjopen-2023-081517

5. Kengia JT, Kalolo A, Barash D, Chwa C, Hayirli TC, Kapologwe NA, et al. Research capacity, motivators and barriers to conducting research among healthcare providers in Tanzania’s public health system: a mixed methods study. Hum Resour Health [Internet]. 2023 Sep 5 [cited 2024 Feb 9];21(1):73. Available from: https://human-resources-health.biomedcentral.com/articles/10.1186/s12960-023-00858-w

6. Van De Goor I, Hämäläinen RM, Syed A, Juel Lau C, Sandu P, Spitters H, et al. Determinants of evidence use in public health policy making: Results from a study across six EU countries. Health Policy [Internet]. 2017 Mar [cited 2024 Feb 26];121(3):273–81. Available from: https://linkinghub.elsevier.com/retrieve/pii/S0168851017300192

7. Van Kammen J, Savigny DD, Sewankambo N. Using knowledge brokering to promote evidence-based policy-making: the need for support structures. Bull World Health Organ. 2006;84(8).

8. Guilbert JJ. The World Health Report 2006: Working together for health [1]. Educ Health Change Learn Pract. 2006;19(3):385–7.

9. Mboera LEG, Rumisha SF, Mbata D, Mremi IR, Lyimo EP, Joachim C. Data utilisation and factors influencing the performance of the health management information system in Tanzania. BMC Health Serv Res. 2021;21(1):4–11.

10. Michie S, van Stralen MM, West R. The behaviour change wheel: A new method for characterising and designing behaviour change interventions. Implement Sci. 2011 Apr 23;6(1).

11. De Leo A, Bayes S, Bloxsome D, Butt J. Exploring the usability of the COM-B model and Theoretical Domains Framework (TDF) to define the helpers of and hindrances to evidence-based practice in midwifery. Implement Sci Commun [Internet]. 2021 Dec [cited 2024 Aug 12];2(1):7. Available from: https://implementationsciencecomms.biomedcentral.com/articles/10.1186/s43058-020-00100-x

12. Kapologwe NA, Kalolo A, Kibusi SM, Chaula Z, Nswilla A, Teuscher T, et al. Understanding the implementation of Direct Health Facility Financing and its effect on health system performance in Tanzania: A non-controlled before and after mixed method study protocol. Health Res Policy Syst. 2019 Jan 30;17(1).

13. United THE, Of R. Comprehensive Council Health Planning Guideline Fifth Edition. 2019;

14. Ministry of Health. Guideline for Developing Comprehensive Hospital Operational Plan (CHOP) for Regional Referral Hospitals. 2016;(August).

15. Mwilike B, Nalwadda G, Kagawa M, Malima K, Mselle L, Horiuchi S. Knowledge of danger signs during pregnancy and subsequent healthcare seeking actions among women in Urban Tanzania: a cross-sectional study. BMC Pregnancy Childbirth [Internet]. 2018 Dec [cited 2024 Sep 21];18(1):4. Available from: https://bmcpregnancychildbirth.biomedcentral.com/articles/10.1186/s12884-017-1628-6

16. Lyimo AA, Guo J, Mushy SE, Mwilike BE. Use of contraceptives and associated factors among male adolescents in rural secondary schools, Coast Region, Tanzania: a school-based cross-sectional study. Contracept Reprod Med [Internet]. 2024 Mar 1 [cited 2024 Sep 21];9(1):8. Available from: https://contraceptionmedicine.biomedcentral.com/articles/10.1186/s40834-024-00268-w

17. Wahed T, Kaukab SST, Saha NC, Khan IA, Khanam F, Chowdhury F, et al. Knowledge of, attitudes toward, and preventive practices relating to cholera and oral cholera vaccine among urban high-risk groups: findings of a cross-sectional study in Dhaka, Bangladesh. BMC Public Health [Internet]. 2013 Dec [cited 2024 Feb 9];13(1):242. Available from: https://bmcpublichealth.biomedcentral.com/articles/10.1186/1471-2458-13-242

18. Subedi D. Explanatory sequential mixed method design as the third research community of knowledge claim. Am J Educ Res. 2016;4(7):570–7.

19. Shiyanbola OO, Rao D. Using an exploratory sequential mixed methods design to adapt an Illness Perception Questionnaire for African Americans with diabetes: the mixed data integration process. 2021;9(1):796–817.

20. Surucu L, Maslakci A. Validity and Reliability in Quantitative Research. Bus Manag Stud Int J. 2020;8(3):2694–726.

21. Jiyenze MK, Tundui C, Sirili N, Mollel H. Effects of implementing a health sector strategic plan: Eidence from Tanzania. East Afr Med J. 2022;99(9):5147–55.

22. Lavis J. Assessing country-level efforts to link research to action. Bull World Health Organ [Internet]. 2006 Aug 1 [cited 2024 Oct 4];84(8):620–8. Available from: https://www.ncbi.nlm.nih.gov/pmc/articles/PMC2627430/pdf/16917649.pdf/

23. Hanney SR, Kanya L, Pokhrel S, Jones TH, Boaz A. How to strengthen a health research system: WHO’s review, whose literature and who is providing leadership? Health Res Policy Syst [Internet]. 2020 Dec [cited 2024 Oct 4];18(1):72. Available from: https://health-policy-systems.biomedcentral.com/articles/10.1186/s12961-020-00581-1

24. Orton L, Lloyd-Williams F, Taylor-Robinson D, O’Flaherty M, Capewell S. The Use of Research Evidence in Public Health Decision Making Processes: Systematic Review. Ross JS, editor. PLoS ONE [Internet]. 2011 Jul 26 [cited 2024 Oct 4];6(7):e21704. Available from: https://dx.plos.org/10.1371/journal.pone.0021704

25. Innvær S, Vist G, Trommald M, Oxman A. Health policy-makers’ perceptions of their use of evidence: a systematic review. J Health Serv Res Policy [Internet]. 2002 Oct 1 [cited 2024 Oct 3];7(4):239–44. Available from: 10.1258/135581902320432778

26. Oliver K, Innvar S, Lorenc T, Woodman J, Thomas J. A systematic review of barriers to and facilitators of the use of evidence by policymakers. BMC Health Serv Res [Internet]. 2014 Dec [cited 2024 Apr 8];14(1):2. Available from: https://bmchealthservres.biomedcentral.com/articles/10.1186/1472-6963-14-2

27. Koon AD. A scoping review of the uses and institutionalisation of knowledge for health policy in low- and middle-income countries. 2020;

28. Lavis JN, Paulsen EJ, Oxman AD, Moynihan R. Evidence-informed health policy 2 – Survey of organizations that support the use of research evidence. Implement Sci. 2008;3(1):1–17.

29. Lavis JN, Wilson MG, Grimshaw JM, Haynes RB, Hanna S, Raina P, et al. Effects of an evidence service on health-system policy makers’ use of research evidence: A protocol for a randomised controlled trial. 2011;

30. Moat KA, Lavis JN. 10 best resources for … evidence-informed health policy making.

31. Gavine A. Maximising the availability and use of high-quality evidence for policymaking: collaborative, targeted and efficient evidence reviews. PALGRAVE Commun. 2018;

32. Armstrong R, Waters E, Dobbins M, Anderson L, Moore L, Petticrew M, et al. Knowledge translation strategies to improve the use of evidence in public health decision making in local government: intervention design and implementation plan. Implement Sci [Internet]. 2013 Dec [cited 2024 Feb 16];8(1):121. Available from: http://implementationscience.biomedcentral.com/articles/10.1186/1748-5908-8-121

33. Armstrong JS. Guidelines for Science: Evidence and Checklists.

34. Smith JD, Hasan M. Quantitative approaches for the evaluation of implementation research studies. Psychiatry Res [Internet]. 2020 Jan [cited 2024 Oct 28];283:112521. Available from: https://linkinghub.elsevier.com/retrieve/pii/S0165178119307024

35. Roberts-Lewis SF, Baxter HA, Mein G, Quirke-McFarlane S, Leggat FJ, Garner HM, et al. The Use of Social Media for Dissemination of Research Evidence to Health and Social Care Practitioners: Protocol for a Systematic Review. JMIR Res Protoc [Internet]. 2023 May 12 [cited 2024 Oct 28];12:e45684. Available from: https://www.researchprotocols.org/2023/1/e45684

36. Brown G, Kyttä M, Reed P. Using community surveys with participatory mapping to monitor comprehensive plan implementation. Landsc Urban Plan [Internet]. 2022 Feb [cited 2024 Oct 28];218:104306. Available from: https://linkinghub.elsevier.com/retrieve/pii/S0169204621002693

37. Sörensen J, Persson AS, Olsson JA. A data management framework for strategic urban planning using blue-green infrastructure. J Environ Manage [Internet]. 2021 Dec [cited 2024 Oct 28];299:113658. Available from: https://linkinghub.elsevier.com/retrieve/pii/S0301479721017205

38. Lim FV, Nguyen TTH. Design-based research approach for teacher learning: a case study from Singapore. ELT J [Internet]. 2022 Oct 14 [cited 2024 Oct 28];76(4):452–64. Available from: https://academic.oup.com/eltj/article/76/4/452/6316630

39. Nabyonga-Orem J, Ssengooba F, Macq J, Criel B. Malaria treatment policy change in Uganda: what role did evidence play? 2014;

40. Hyder F, Rothman DL. Evidence for the importance of measuring total brain activity in neuroimaging.

41. Bennett NJ. Using perceptions as evidence to improve conservation and environmental management. Conserv Biol. 2016;30(3).

42. Hollingworth SA. What do we need to know? Data sources to support evidence-based decisions using health technology assessment in Ghana. . Epidemiology. 2020;

